# Combining new interventions with urban development as a path to effective, consistent, and durable control of dengue

**DOI:** 10.64898/2026.03.11.26348180

**Authors:** T. Alex Perkins, Katie M. Susong, Katie Tiley, Abhijit Majumder, Saikanth Ratnavale, Manar Alkuzweny, Moritz U.G. Kraemer, Hannah E. Clapham, Oliver J. Brady

## Abstract

Dengue is a mosquito-transmitted viral disease that has long defied control but now has a growing list of promising interventions becoming available. Major investments in these interventions are being made without a clear picture of the long-term implications of those choices. We used a mathematical model to project the impacts of alternative suites of interventions across 1,634 cities to the year 2050. We found that recently developed interventions have the potential for significant reductions in dengue hospitalisations over a period of a few years. Beyond that timeframe, our model predicts a buildup of susceptibility that diminishes intervention effectiveness over time. Routine vaccination leads to only modest reductions when layered on top of other interventions. The most effective and sustainable strategy combines short-term investments in new interventions with long-lasting changes to remove mosquito habitat in urban environments, resulting in *>* 90% reductions in disease burden in all cities over a 25-year period.

## Introduction

The current time is a vexing one for those seeking to control dengue, the world’s most medically significant mosquito-transmitted viral disease. On the one hand, new interventions are becoming available that arguably show greater promise than any approach since Soper’s 1960s campaign to eradicate the *Aedes aegypti* mosquitoes that transmit dengue virus (DENV). The most promising interventions today include vaccines (specifically, Qdenga by Takeda) [1] and *Wolbachia* replacement (specifically *wMel Wolbachia*, an endosymbiotic bacterium that reduces mosquito infectiousness) [2]. On the other hand, reported dengue cases have broken records year over year for much of the past two decades. In 2024, 14.1 million cases were reported globally, double the 7 million reported the year before [3].

This juxtaposition of dengue’s seemingly never-ending rise with the emergence of promising new interventions makes the future of dengue exceptionally uncertain. Moreover, with increasingly strained public health resources in settings where dengue is a problem [4], officials are being forced to make choices about which new interventions to invest in, if any. While arguments can be made about the relative merits of dengue vaccines, *Wolbachia*, and enhanced vector control, available evidence is generally limited to short-term data from specific contexts on the isolated, rather than combined, effects of these interventions [1, 2]. These limitations point to a role for mathematical modeling in generating much needed insight about how these interventions can be used to greatest effect.

Recently, there has been increasing acknowledgment that, in addition to more interventions, sustainable dengue control will also need to address the root causes of why invasive *Aedes* mosquitoes have been able to colonize urban environments in the first place [5]. “Building out *Aedes*” is an approach that promotes combining intensive use of new and existing dengue control interventions with longer-term changes to the way urban environments are built and managed to reduce the habitability of cities for mosquitoes. How these two components (reducing transmission and reducing the potential for transmission) interact over time is important for designing an effective and sustainable building out *Aedes* strategy. Our goal is to assess the interaction of these short and long-term interventions with mathematical models.

Gaining robust insights from mathematical models about dengue control is not straight-forward, however. One complicating factor for modeling is the vast extent and variability in DENV epidemiology globally. More than 5 billion people spanning 169 countries live in (mostly urban) areas at risk of DENV transmission [6]. Those settings vary in their intensity of transmission, the seasonality of transmission, demographic attributes, health infrastructure, and in many other respects. Despite that, mechanistic models of dengue prevention and control are typically designed to consider either a generic setting, one specific setting, or, at best, a small number of representative settings [7, 8, 9, 10].

While mechanistic models have generally eschewed geographic variability in dengue epidemiology, a growing number of resources have become available in recent years that provide specific, quantitative insights into different layers of dengue’s epidemiology globally. Two key resources include high-resolution global maps of dengue virus force of infection [11] and the occurrence probabilities of the mosquito species that transmit it [12]. High-resolution temperature estimates, when combined with laboratory-based estimates of the effects of temperature on mosquito and virus traits [13], offer additional insights. Furthermore, robust estimates of the effectiveness of these new interventions are now available across a growing number of geographic settings [14, 15, 5]. In this study, we leveraged those resources to develop a mechanistic model of DENV transmission and control in every major city at risk of DENV transmission worldwide, which we used to obtain insights about the promise of alternative strategies for the control of dengue across a wide range of transmission settings.

## Results

### Validation against epidemiological data

We found that our model’s predictions of seasonal patterns of dengue transmission were consistent with independent temporal data on dengue incidence from 803 cities where such data were available. In terms of the timing of seasonal highs and lows, we found that our model predicted the high month of dengue incidence within two months of the observed value in 609/803 cities (75.8%) (Fig. 1A) and the low month of dengue incidence within two months of the observed value in 668/803 cities (83.2%) (Fig. 1B). The overall correlation of predicted versus observed monthly dengue incidence was positive in 698/803 cities (86.9%) and greater than 0.5 in 503/803 cities (62.6%) (Fig. 1C). One metric of seasonality in which the model underperformed was epidemic intensity [16], with the model predicting a more pronounced seasonal epidemic pattern than the incidence data suggested (Fig. 1D). Even so, the model performed reasonably well at predicting which cities had relatively more intense seasonal epidemics than others (Kendall’s tau: 0.34). As expected, the time-averaged dengue virus force of infection from our model matched well with the values from Cattarino et al. to which it was calibrated (Pearson correlation: 0.997) (Fig. S1), meaning that our model is consistent with current understanding of spatial variation in dengue transmission.

**Figure 1:**
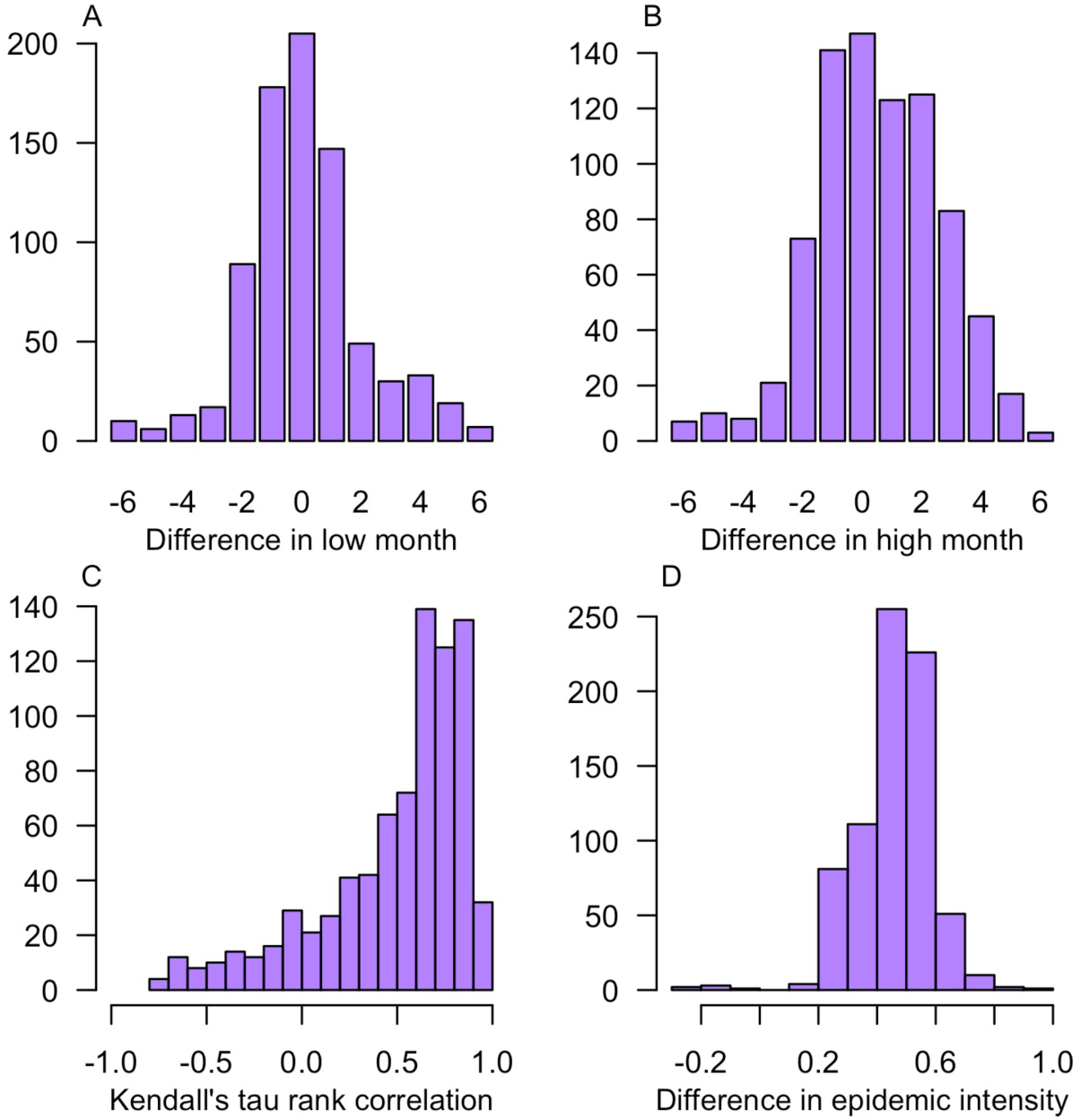
Validation of the model’s predictions of seasonal patterns. We compared model predictions of infection prevalence aggregated monthly against monthly dengue case reports from OpenDengue for 803 cities where data for all 12 months were available. Consistency between model predictions and those independent data were assessed on the basis of four metrics (A-D).

### Model predictions of seasonal variation in transmission potential

Our model’s predictions of seasonal dengue incidence patterns are driven by predictions of seasonal variation in transmission potential, as captured by the time-varying basic reproduction number, *R*_0_(*t*). These predictions vary considerably across cities, in part due to latitudinal variation in the timing of summer (Fig. 2A). The number of days per year when *R*_0_(*t*) exceeds 1 is also strongly dependent on latitude, with cities farther from the equator having fewer days per year that are suitable for transmission (Pearson correlation: -0.712) (Fig. S2). When population susceptibility is taken into account, the time-varying effective reproduction number, *R*(*t*), follows nearly identical seasonal patterns as *R*_0_(*t*) but with lower values (Fig. 2B). This results in no cities having year-round transmission potential when population susceptibility is taken into account (i.e., no cities where *R*(*t*) *>* 1 for all *t*) (Fig. 2C). Among cities that do not have year-round transmission potential, the median number of days with a value greater than 1 is 174 for *R*_0_(*t*) (inter-quartile range [IQR]: 122–226) and 125 for *R*(*t*) (IQR: 101–162) (Fig. 2C), meaning that cities are capable of supporting local dengue transmission for around four months per year, on average.

**Figure 2:**
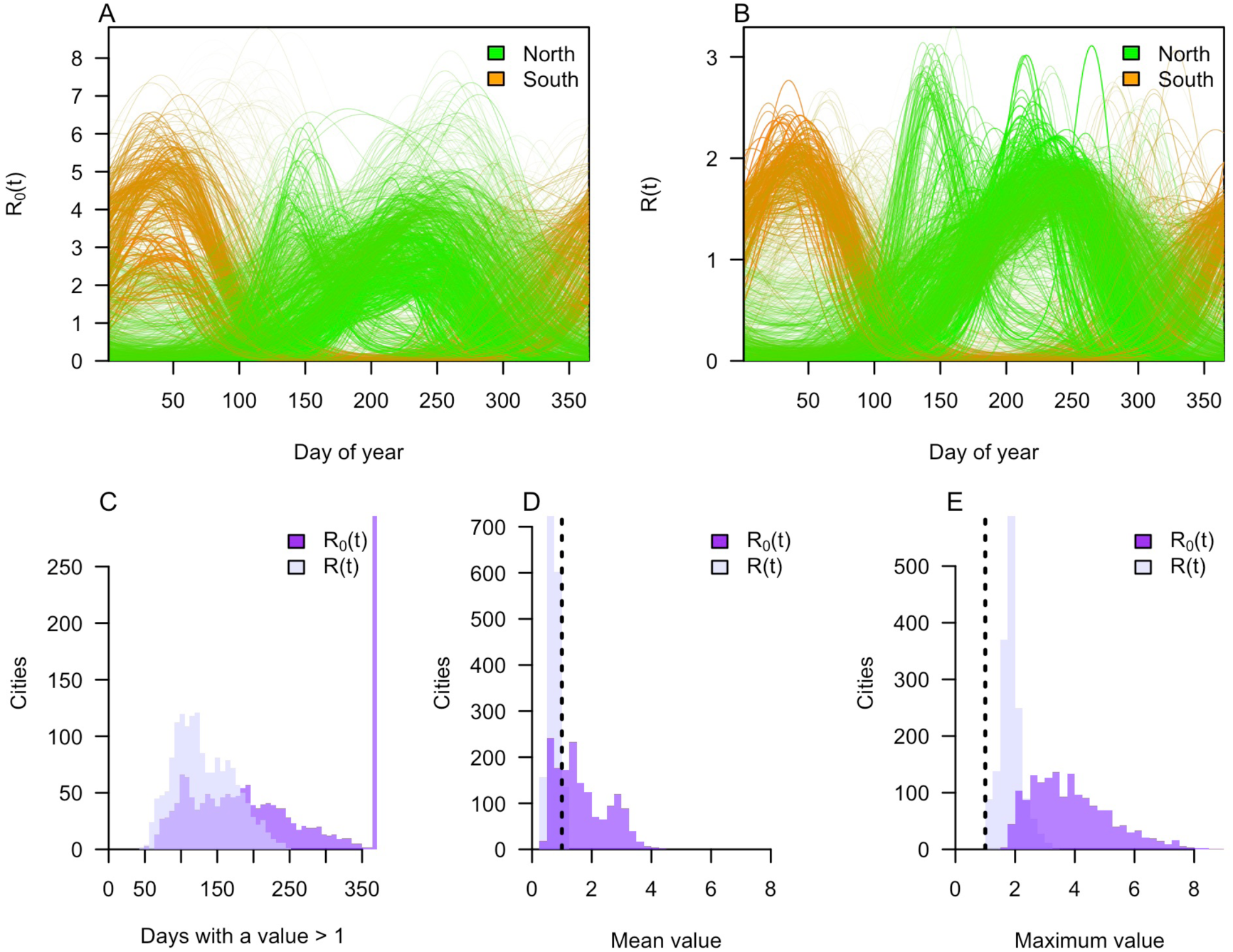
City-specific predictions of seasonal transmission. For each of 1,634 cities, the model predicted seasonally varying values of the basic reproduction number, *R*_0_(*t*), (A) and the effective reproduction number, *R*(*t*) (B). Cities with extreme north (red) and south (blue) latitudes are emphasized with thicker, darker lines to illustrate correspondence between transmission seasons and the timing of summer in each hemisphere. Maximum (C) and median (D) values of *R*_0_(*t*) and *R*(*t*), as well as the number of days they exceed 1 (E), are also shown.

In addition to variation in timing, the model also predicted considerable variation in the intensity of transmission. For *R*_0_(*t*), 95% of cities had seasonal mean values between 0.54 and 3.42 (Fig. 2D) and seasonal maximum values between 2.0 and 6.99 (Fig. 2E). With a median of 3.67 for the seasonal maximum value of *R*_0_(*t*), an intervention would need to reduce transmission by nearly 75% (following 1 *→* 1*/R*_0_) to eliminate local dengue transmission in half of the cities we examined, in the absence of population immunity. An intervention that reduces transmission by 50% would achieve this in only 2.5% of cities. For *R*(*t*), the 95% quantile ranges of the seasonal average (0.44–1.0) (Fig. 2D) and seasonal maximum (1.14–2.75) (Fig. 2E) values were more moderate. Thus, in the presence of current levels of population immunity, a 50% reduction in transmission would reduce *R*(*t*) below 1 year-round in more than half of cities, and a 75% reduction would have this effect in all cities.

### Exploring how the dynamic feedback of immunity undermines long-term effectiveness in a representative city

While seasonal maximum values of *R*_0_(*t*) and *R*(*t*) provide convenient rules of thumb about requirements for control, they either overestimate (in the case of *R*_0_(*t*)) or underestimate (in the case of *R*(*t*)) control requirements over time. Initially, reducing transmission by a factor 1 *→* 1*/* max*{R*(*t*)*}* would be sufficient to control dengue. Over time, susceptibility would grow and the degree to which transmission would need to be reduced would approach 1 *→* 1*/* max*{R*_0_(*t*)*}*. The timing of how this unfolds depends on a complex interplay between demographic turnover and transmission dynamics in a way that is highly context-dependent.

To understand how requirements for control change over time, we applied optimal control theory to determine an optimal schedule for the application of enhanced vector control in each of the 1,634 cities in our analysis. For an illustrative example, we focus on São Paulo, Brazil, which has a seasonal average value of *R*_0_ of 1.41, near the global median of 1.45, and where our model successfully predicted dengue seasonality (peak model-predicted force of infection in March compared with peak reported hospitalisations in March and April). Our default assumptions about enhanced vector control are that it can reach a maximum of 30% coverage and that a single application reduces mosquito density by 97% in treated spaces, that the mosquito population recovers after 25 days, and that a single application costs $0.0846 per person. Applications may occur repeatedly as long as their cost is offset by reduced costs of dengue illness.

Under these assumptions, our model predicted that transmission could be reduced substantially for several years following the introduction of enhanced vector control (Fig. 3A). This required continuous applications for two months in the first year of enhanced vector control (Fig. 3B). During this period of continual application, max*{R*(*t*)*}* was reduced from 1.96 to 1.56 (Fig. 3C). After ten years of enhanced vector control, seasonally averaged population susceptibility rose from its historical value of 0.38 up to 0.47 (Fig. 3D), resulting in an increase in max*{R*(*t*)*}* to 1.88 (Fig. 3C). Meanwhile, the required control effort rose to four months of continuous application by the tenth year (Fig. 3B). After 25 years of enahnced vector control, our model predicted a “new normal” would be reached that involves moderately reduced transmission compared to historical patterns, albeit with a new dependency on more than twice as much enhanced vector control as required initially. Failure to keep up with required control at any point beyond then would lead to a major outbreak, given the significant increase in susceptibility over that 25-year period of enhanced vector control.

**Figure 3:**
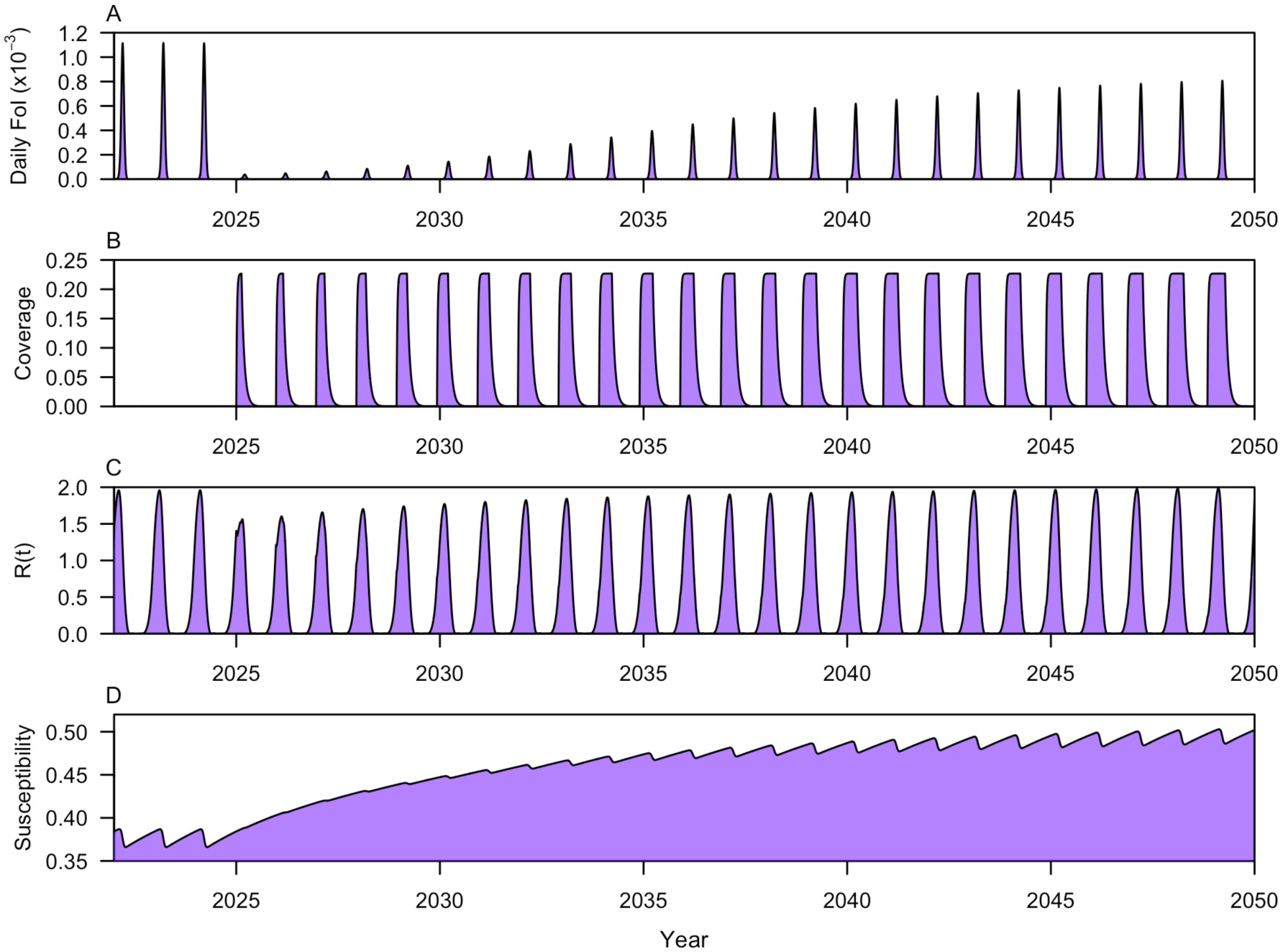
Example of the model’s dynamics in São Paulo, Brazil. To illustrate the behavior of key variables, we simulated the model under baseline conditions through 2024 and then allowed for enhanced vector control beginning in 2025. This resulted in a significant reduction in the force of infection for several years followed by a steady increase after (A). This increase in force of infection occurred despite an increase over time in the proportion of the year in which control was applied, as determined by optimal control theory (B). The initial introduction of control resulted in a decrease in *R*(*t*) at first (C), but this was later counteracted by an increase in susceptibility over time following the initial success of enhanced control (D).

### Variability in impacts and costs across cities

Even though enhanced vector control resulted in diminished impacts at greater costs over time in our illustrative example, a 58% reduction in dengue hospitalisations was achieved over a 25-year period, which is substantial. To understand how variable such impacts might be across cities, we expanded on our illustrative example to quantify cumulative impacts and costs over a 25-year period across 1,634 cities.

Under the scenario of enhanced vector control, city-level reductions in hospitalisations over a 25-year time frame ranged from near zero to 98% (Fig. 4A). This variation among cities was strongly negatively associated with several measures of transmission intensity (Fig. S3), which is a consequence of the greater difficulty of control in settings with high transmission. In contrast, baseline population susceptibility to dengue was positively associated with proportional reduction in hospitalisations, which is a consequence of susceptibility being a decreasing function of transmission intensity in the model (Pearson correlation: -0.91). Similarly, birth rate was positively associated with proportional reduction in hospitalisations, due to an inverse relationship between birth rate and force of infection, which are independent inputs to the model (Pearson correlation: -0.16) (Fig. S3). At a given transmission intensity, a higher birth rate would result in a faster increase in susceptibility following initial success with control and, therefore, greater difficulty in reducing transmission.

**Figure 4:**
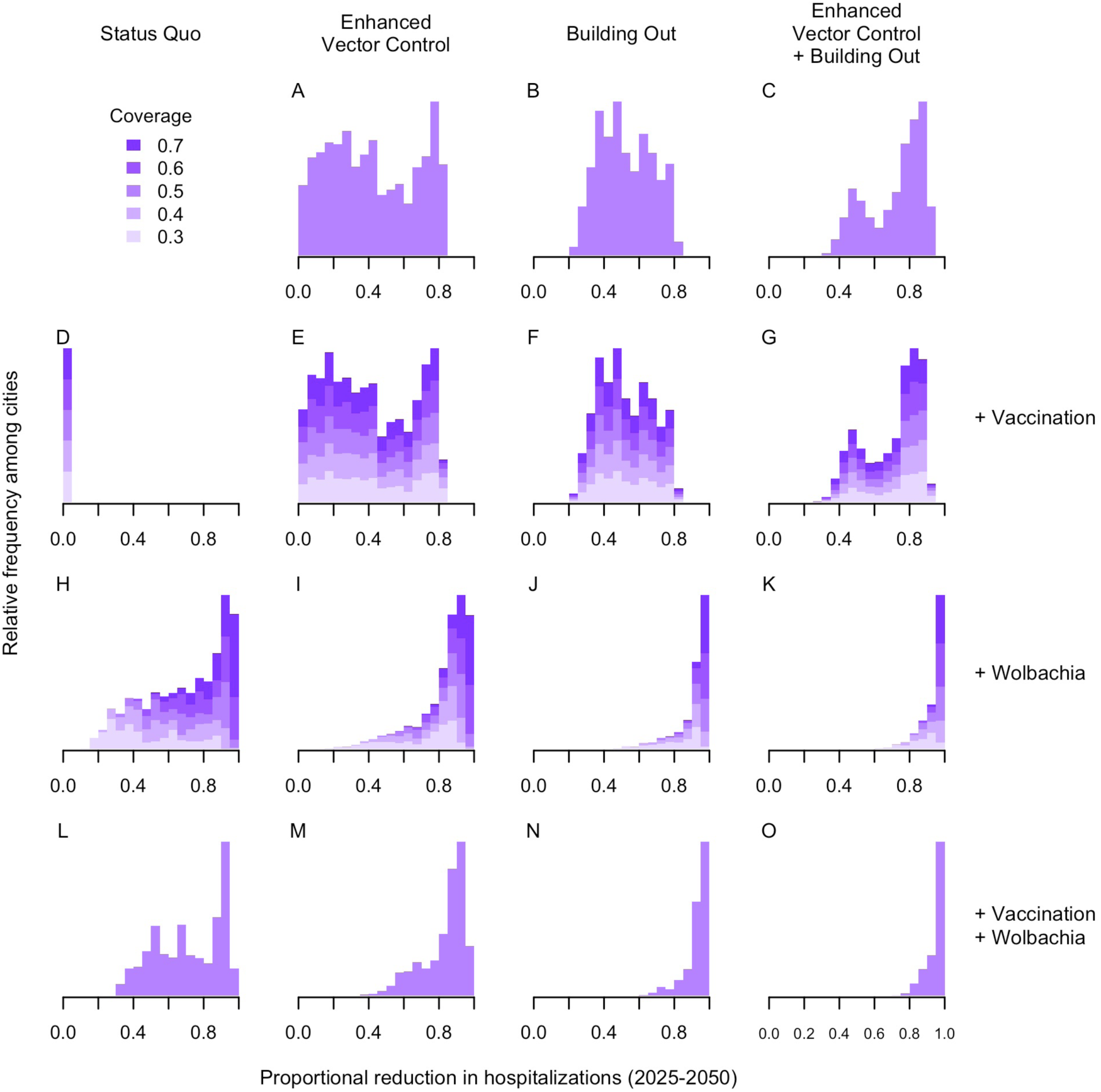
Histograms of predicted proportional reductions in cities under scenarios of different combinations of interventions. Proportional reduction is calculated as cumulative hospitalizations over a 25-year time period compared to a baseline scenario of sustained current (non-enhanced) control efforts with higher values indicating larger reductions in cases. “Building out” describes a scenario where gradual improvements to the urban environment remove habitat for *Aedes* mosquitoes.

We then expanded our simulation to include the novel interventions of vaccination, *Wolbachia* replacement, and “Building out *Aedes*”—a program of gradual improvements to the building and management of urban environments to remove egg-laying habitats from cities. The predicted reduction across cities in the *Wolbachia*-only scenario was also variable and dependent on the level of coverage that can be achieved, a key uncertainty following recent variable coverage levels observed in different release sites worldwide [17]. Under a relatively pessimistic scenario of a maximum *Wolbachia* coverage of 30%, only 6.5% of cities are predicted to achieve a reduction in hospitalisations of 85% or more, whereas if coverage can be increased to 70% (closer to the levels achieved in trials [14]) this increases to 72% of cities (Fig. 4H). In contrast, under a scenario of vaccination only and at the highest vaccination coverage we considered, the range of percent reductions in dengue burden was limited to no more than 2% (Fig. 4D). This more muted impact of vaccination reflects our assumption of no indirect effects (i.e. no reduction in DENV transmission, only reduced disease), which make these results a lower bound on the possible impacts of vaccination.

In general, combinations of two or more interventions performed more consistently well across cities than single interventions. For example, even though their impacts alone were similar, combining enhanced vector control with *Wolbachia* replacement shifted the overwhelming majority (65% - 99% depending on coverage) of cities to reductions in hospitalizations of 85% or more, even if only 30% penetrance of *Wolbachia* can be achieved (Fig. 4). This synergy can be attributed to the fact that the combination of these interventions reduces *R*_0_(*t*) by such an extent that rises in susceptibility are not sufficient to push *R*(*t*) above 1, provided that enhanced vector control continues to be used. As before, variability in the extent of these impacts across cities was negatively associated with transmission intensity (Fig. S4).

To assess sustainability, we projected cumulative annual costs over the 25 year time period and compared annual costs in the first and last years of the program (Fig. 5). While enhanced vector control has lower initial costs, over time these costs accumulate and increase proportional to need as population susceptibility increases, with annual costs increasing by an average of 116% over a 25-year period. Investing in new vaccination or *Wolbachia* replacement programs alone has higher costs initially but entails lower costs in later years of the program. Cumulatively, a combined strategy of vaccination and enhanced vector control is the most costly over 25 years (median: $12.39 per person; 95% range: $7.76-21.23), given that vaccination is relatively costly and similar levels of enhanced vector control are required regardless of vaccination. In contrast, *Wolbachia* replacement reduces the cumulative cost of enhanced vector control from a median of $3.16 per person over 25 years (95% range: $1.53-5.52) to $1.73 ($0.08-3.85). This savings comes at the cost of the initial investment in *Wolbachia*, but also comes with the benefit of a greatly enhanced reduction in hospitalisations (Fig. 4A vs. 4I), particularly in settings with higher transmission.

**Figure 5:**
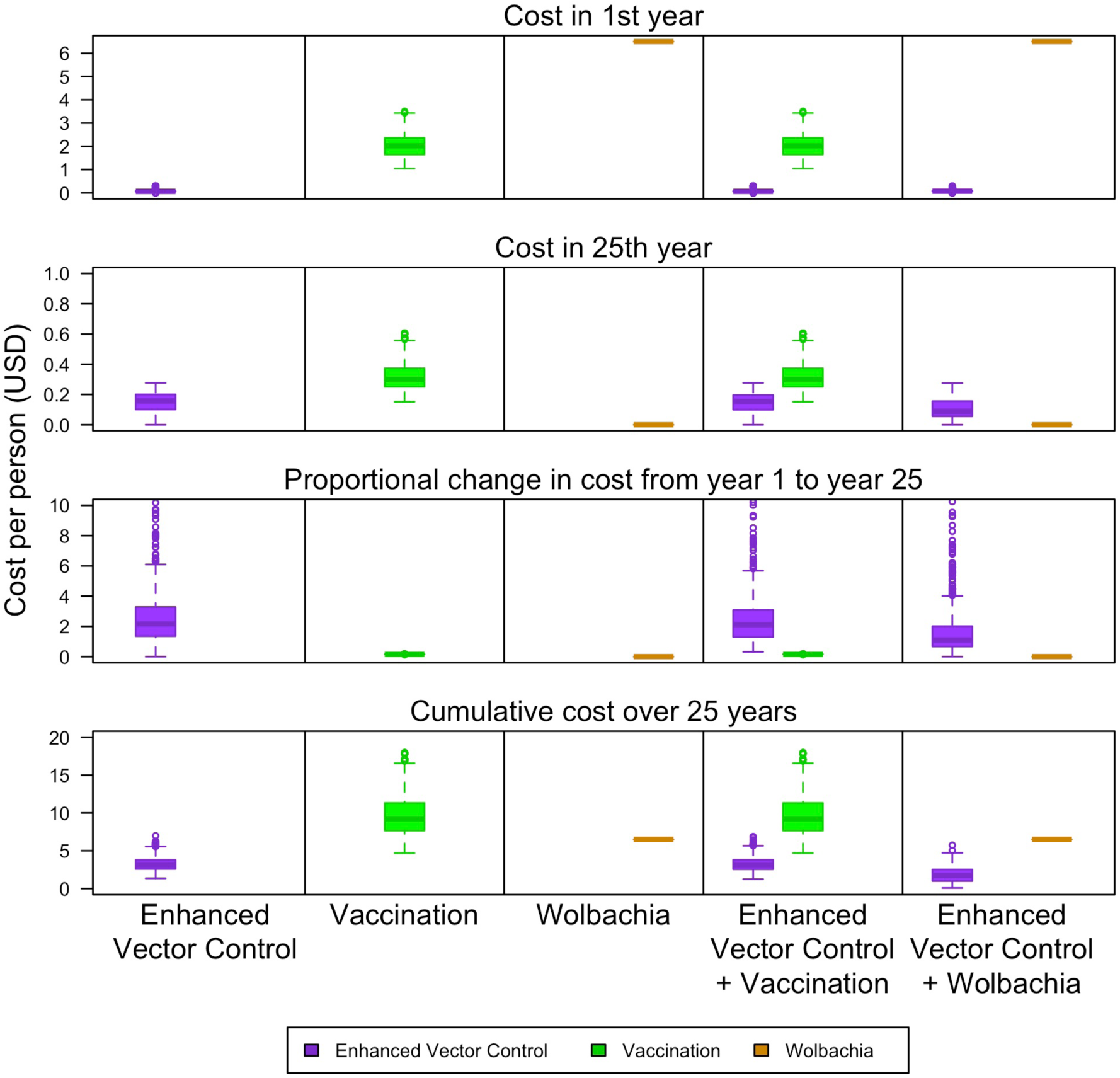
Costs of alternative control strategies. Under scenarios of enhanced vector control, vaccination, and *Wolbachia* alone, as well as enhanced vector control combined with each of the other two (columns), we calculated the costs associated with each intervention (colors) as a distribution across the 1,634 cities in our analysis. Costs are presented within the first year only, within the 25th year, as a proportional change between those two costs, and cumulatively over the 25-year period (rows). All costs are defined on a per person basis, meaning that city-wide costs are divided among all residents of the city rather than being divided among those who are treated.

To contemplate how the high ongoing costs of the aforementioned interventions could be counteracted, we explored how the impacts of a gradual but persistent decline in *R*_0_ resulting from building out *Aedes* (modeled after Singapore) would change predictions about intervention impact. On its own, Building out Aedes had a similar impact as enhanced vector control alone (Fig. 4A vs. 4B), due to the fact that it took a number of years for its impacts to register. However, when building out was combined with enhanced vector control (Fig. 4C) or *Wolbachia* (Fig. 4J), 86-94% of cities saw reductions in dengue burden of 80% or more over a 25-year time horizon. When all three interventions were combined (Fig. 4K), 99.9% of cities saw this level of reduction. These synergies arise from the complementary timing of strategies whose impact is most acute in the short term (enhanced vector control, *Wolbachia*) and long term (building out). If building out *Aedes* can be achieved as part of a program of ongoing urban renewal or with relatively modest one-time investment costs comparable to those associated with *Wolbachia*, then the strategy of combining building out *Aedes* with enhanced vector control and/or novel control tools could result in the most sustainable and widely applicable strategy for dengue control worldwide.

## Discussion

Here, we present a new modelling framework for exploring the long-term effectiveness of different combinations of novel interventions against dengue and their variability in different cities around the world. Previous modelling work has raised the possibility of novel interventions leading to short-term reductions in dengue followed by long-term resurgence [18, 19, 20]; our work grounds those theoretical concerns within a realistic depiction of the spatiotemporal variability of DENV transmission and in the context of recent developments around leading interventions. Specifically, we show that combining interventions that lead to immediate reductions in transmission (enhanced vector control and *Wolbachia* replacement) with long-term changes to remove mosquito habitat (Building out *Aedes*) will lead to the largest, most certain, and most sustainable long-term burden reductions. Our results show that this combination of short- and long-term interventions is likely to be a necessary approach for many dengue-endemic cities across the tropics, especially those with higher transmission.

Importantly, our work shows that achieving specific public health objectives may be possible through different combinations of interventions, provided that such combinations include a complimentary mix of short-term and long-term effects. For example, cities that struggle to achieve high penetrance of *Wolbachia* [17] or face community acceptance issues [21] can alternatively invest in enhanced vector control, or vice versa in the case of cities with high levels of insecticide resistance [22] or other barriers to enhanced vector control. Similarly, different cities will undoubtedly require different paths to realise the long-term benefits associated with the Building out *Aedes* paradigm. A range of locally appropriate examples are discussed at length elsewhere [5].

Our results also suggest that the long-term effectiveness of novel interventions may be difficult to predict based solely on the reduction in reported cases in the first few years of evaluation. In agreement with other *Wolbachia* modelling studies [23], we predict that, even at low efficacy levels, the initial decline in transmission in many cities could be substantial due to high levels of naturally acquired immunity. Once this immunity erodes due to demographic turnover, our model predicts that in many settings *Wolbachia* replacement will be insufficient to prevent resurgence, as has been observed with vector control in Singapore [24]. Depending on the setting, it may be many years before the difference between high- and low-effectiveness novel interventions becomes measurable, thus cautioning against relying on case counts from routine surveillance as the only method of monitoring long-term effectiveness. This also has implications for the current WHO recommendation [25] to target vaccines and *Wolbachia* to high burden settings, where rebound is most likely. Routinely measuring seroprevalence can measure long-term changes in populaiton immunity and changes in FOI.

More broadly, this work shows the value of integrating dynamic models with geospatial maps. Previous model-based estimates of *Wolbachia* or vaccination impact have restricted dynamic model simulations to a handful of generalizable or highly aggregated settings [7, 8, 9, 10] that underrepresent the variety and heterogeneity in transmission globally [26]. Alternatively, static estimates of intervention impact using maps [27, 11] neglect the importance of intra- and inter-annual variations in transmission and non-linear feedback between interventions, transmission, and immunity. Our modelling framework combines the strengths of both approaches and could be used to explore other interventions, or to identify new climatic or immunity-related drivers of global dengue dynamics.

Our findings also show the growing importance of more targeted use of enhanced vector control interventions. Our results suggest that novel interventions can reduce the annual average reproduction number below 1 in many settings, but climate variation and less frequent use of vector control will cause temporary periods where the reproduction number exceeds 1, allowing outbreaks to occur. Strong environmental, entomological, and epidemiological surveillance will be needed to identify areas and times at risk so additional vector control can be appropriately targeted. There is a consensus that adoption of novel interventions will still need to be supported by continued vector control, but little consideration has been given to how surveillance and response capacity will need to change to ensure a sustainable integrated program.

To a certain extent, these findings will depend on our limited assumptions about dengue immunity. In assuming even circulation of serotypes and lifelong immunity, our models align with common practice in the dengue modelling field [28], but complexities such as partial cross immunity and serotype cycling [29] have been documented and could lead to faster rebound times and lower long-term effectiveness of strategies that only achieve partial control. The lack of inter-annual variability is also a limitation as high transmission years driven by multi-year trends (e.g., El Niño [30]) may lead to large outbreaks in settings where control is more feasible in a long-term average year. We also did not account for future changes in climate, urbanisation or travel, all of which are expected to increase transmission intensity and increase the need for scenarios that use combinations of interventions. Our assumption that dengue vaccines only prevent disease and not transmission is admittedly conservative and leads to less favourable estimates of vaccination impact when compared to other interventions. At the same time, this conservatism is balanced by our optimistic assumption of no waning in protection (cf. [9, 31]). Finally, we caution city-level stakeholders against relying exclusively on these global estimates of long-term effectiveness for decisions in any single city. Dengue virus serotype dynamics, inter-annual weather variation, and undocumented changes in surveillance practices over time all contribute to sub-optimal fidelity of our model in various cities. While the predictions made here about the relative impact of different intervention strategies are unlikely to be affected, estimates of cost and long-term effectiveness may differ and should be refined with local modelling studies tailored to specific city contexts [32].

The next step will be for national and city-level policymakers to work with modellers to adapt modelling frameworks, such as this, to better reflect the local epidemiology of their settings and identify reasonable long-term policy options for arbovirus control. Predictions from these models can support investment cases for novel interventions [33] or expansion of municipal services to “build out *Aedes* mosquitoes” that will ultimately lead to more cost-effective, sustainable reductions in burden [5].

## Methods

### Overview of the model

We developed a dynamic transmission model of dengue virus (DENV) to simulate long-term transmission dynamics and evaluate the impact of multiple interventions—both alone and in combination—across over 1,600 cities globally. The model is age-structured and serotype-aggregated, with immunity tracked over successive DENV infections. It also accommodates different intervention modalities, including enhanced vector control, vaccination, *Wolbachia*, and a suite of enhancements of urban infrastructure dubbed “Building out Aedes” [5].

The model tracks infection status across twelve states in total, including susceptible, infectious, and recovered states for individuals experiencing their first, second, third, or fourth DENV infection. With the exception of those experiencing their fourth and final serotype infection, individuals remain in the recovered state and protected against all serotypes for a year before becoming susceptible again. Individuals with *j* previous serotype infections experience a reduction in force of infection by a factor 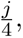 as well as increasing rates of background mortality given that individuals with more previous infections tend to be older. To maintain overall population size, births occur at a rate that offsets deaths and contributes to the fully susceptible class. The number of previous serotype infections also determines the probability that one of four infection outcomes—non-medical, ambulatory, hospitalized, or fatal—is experienced. These infection outcomes have no bearing on transmission dynamics but do impact the calculation of disease-associated costs.

Mosquitoes are not incorporated into the model explicitly, but mosquito and virus parameters do shape the time-varying transmission coefficient. Mosquito and virus parameters were combined mathematically according to standard reasoning about their roles in the transmission cycle [34], including the mosquito-human ratio, the mosquito blood-feeding rate, the probabilities of human-mosquito and mosquito-human transmission, the duration of human infectiousness, the adult mosquito mortality rate, and the incubation period of the virus in the mosquito. Several of these parameters are influenced by temperature and other climate factors that are primary drivers of temporal variability in the model.

The coverage of enhanced vector control is treated dynamically with its own state variable. This coverage wanes as the time-limited effectiveness wanes, and it waxes as enhanced vector control is newly applied or reapplied. The portion of a city where enhanced vector control was applied recently enough to still be effective experiences a temporary reduction in the mosquito-human ratio. To narrow the range of possibilities for the application rate, we applied optimal control theory to identify a time-varying application rate that minimized the combined cost of enhanced vector control and direct costs of disease cumulatively over time. This optimization can be rationalized as resulting from mosquito control practitioners applying either early warning systems or local knowledge of the seasonal timing of transmission in their city, which is key for developing a model that is suitable for the varied geographies to which we applied this model.

The coverage patterns of the other interventions in the model unfold according to pre-specified trajectories given that they entail long-term investments and have greater permanency than enhanced vector control. In the case of vaccination and *Wolbachia*, their coverage increases over a five-year period, after which their maximum coverages are maintained. In the case of building out, coverage increases linearly for every year of the simulation. *Wolbachia* and building out both result in a proportional reduction in the transmission coefficient, whereas vaccination has no impact on transmission but reduces the severity of infection outcomes among vaccine recipients. Empirical data suggest that any impact on transmission conferred by Qdenga is likely to be modest and short-lived [35], making the latter assumption conservative but reasonable.

### Tailoring the model to different cities

We implemented the model independently for all cities in areas with a predicted forece of infection (FOI) ¿0 [11] and with over 100,000 residents (n = 1,634 cities) [36], spanning a wide range of epidemiological, demographic, and economic settings. Seasonality in the time-varying transmission rate is driven in large part by seasonal temperature variation. We used satellite-derived ERA5 re-analysis estimates of daily minimum and maximum temperatures [37] to inform diurnally and seasonally varying values of four temperature-sensitive mosquito and virus parameters specific to each city to which we applied our model. In addition, we used modelled estimates of *Aedes aegypti* occurrence probability on a monthly basis [38] to derive a proxy of the mosquito-human ratio specific to each city and month. While these parameters are capable of generating a biologically plausible pattern of relative transmission, they require additional calibration to reproduce the magnitude of transmission. To accomplish this, we tuned a parameter for each city that resulted in a multi-year average of the dengue virus FOI matching modeled estimates of this quantity produced by Cattarino et al. [11].

While the aforementioned transmission parameters were modeled at a city level due to the high spatial resolution of their inputs, other aspects of the model were specified at a national level. These include demographic inputs of age-specific mortality and age distribution [36]. While the model has the potential to incorporate country-specific information about the costs of dengue illness and costs of interventions, we chose to standardize all costs to a single country (Brazil) to emphasize differences in results attributable to epidemiological, rather than economic, factors. Intervention effectiveness was parameterized at a global level based on prominent national and international randomized controlled trials (Supplementary Methods intervention parameters section). The costs of dengue illness (direct and indirect) were taken from a previous global estimation study [39] inflation-adjusted to 2025 values while the costs of interventions were based on reported cost estimates from recent large-scale programmes (Supplementary Methods intervention parameters and cost of disease parameters sections).

### Validation against epidemiological data

Through its construction, our model is calibrated to global estimates of spatial variation in dengue transmission intensity, which were themselves validated against seroprevalence surveys as part of their original publication [11]. The primary aspect of the model that does require validation are its predictions about seasonal variability in transmission, which reflect a plausible, but unvalidated, set of assumptions. To address this, we compared the model’s predictions to data on relative monthly dengue incidence (proportion of annual cases each month) reported by public health surveillance systems and collated by the OpenDengue project [40, 41]. These data were not used at any stage of model parameterization and, therefore, constitute an independent basis for model validation.

### Model outputs and intervention scenarios

Because mortality risk for dengue is relatively low and its greatest burden occurs in the form of hospitalizations, we focused on cumulative hospitalizations as our primary epidemiological output of interest. Our primary economic outputs of interest were the additional costs of scaled up interventions. We considered a total of 38 intervention scenarios, reflecting different combinations of enhanced vector control, vaccination, *Wolbachia*, and building out, as well as varying maximum coverages of *Wolbachia* replacement and vaccination. All simulations were carried out with the introduction of interventions beginning in 2025 and ending in 2050 assuming no changes in environmental variables.

## Data Availability

All data produced in the present study are available upon reasonable request to the authors.

## Acknowledgements

We thank the University of Notre Dame Center for Research Computing for computing resources.

## References

[1] Wilder-Smith, A., Cherian, T. & Hombach, J. Dengue Vaccine Development and Deployment into Routine Immunization. Vaccines 13, 483 (2025). URL https://www.mdpi.com/2076-393X/13/5/483.

[2] Paz-Bailey, G., Jernigan, D. B., Laserson, K., Zielinski-Gutierrez, E. & Petersen, L. New solutions against the dengue global threat: opportunities for Wolbachia interventions. International Journal of Infectious Diseases 157, 107923 (2025). URL https://linkinghub.elsevier.com/retrieve/pii/S1201971225001468.

[3] Haider, N. et al. Global dengue epidemic worsens with record 14 million cases and 9000 deaths reported in 2024. International Journal of Infectious Diseases 158, 107940 (2025). URL https://linkinghub.elsevier.com/retrieve/pii/S120197122500164X.

[4] Malavige, G. N., et al. Facing the escalating burden of dengue: Challenges and perspectives. PLOS Global Public Health 3, e0002598 (2023). URL https://dx.plos.org/10.1371/journal.pgph.0002598.

[5] Brady, O. et al. Building out Aedes mosquitoes: Eliminating dengue as an urban threat. Lancet **in review** (2026).

[6] Bhatt, S. et al. The global distribution and burden of dengue. Nature 496, 504–507 (2013).

[7] Flasche, S., et al. The Long-Term Safety, Public Health Impact, and Cost-Effectiveness of Routine Vaccination with a Recombinant, Live-Attenuated Dengue Vaccine (Dengvaxia): A Model Comparison Study. PLOS Medicine 13, e1002181 (2016). URL https://dx.plos.org/10.1371/journal.pmed.1002181.

[8] Hladish, T. J. et al. Designing effective control of dengue with combined interventions. Proceedings of the National Academy of Sciences 117, 3319–3325 (2020). URL https://pnas.org/doi/full/10.1073/pnas.1903496117.

[9] Cracknell Daniels, B., Ferguson, N. M. & Dorigatti, I. Efficacy, public health impact and optimal use of the Takeda dengue vaccine. Nature Medicine 31, 2663–2672 (2025).

[10] Shen, J., et al. Vaccination strategies, public health impact and cost-effectiveness of dengue vaccine TAK-003: A modeling case study in Thailand. PLOS Medicine 22, e1004631 (2025). URL https://dx.plos.org/10.1371/journal.pmed.1004631.

[11] Cattarino, L., Rodriguez-Barraquer, I., Imai, N., Cummings, D. A. T. & Ferguson, N. M. Mapping global variation in dengue transmission intensity. Science Translational Medicine 12, eaax4144 (2020). URL https://www.science.org/doi/10.1126/scitranslmed.aax4144.

[12] Kraemer, M. U., et al. The global distribution of the arbovirus vectors Aedes aegypti and Ae. albopictus. eLife 4, e08347 (2015). URL https://elifesciences.org/articles/08347.

[13] Mordecai, E. A., et al. Detecting the impact of temperature on transmission of Zika, dengue, and chikungunya using mechanistic models. PLOS Neglected Tropical Diseases 11, e0005568 (2017). URL https://dx.plos.org/10.1371/journal.pntd.0005568.

[14] Utarini, A. et al. Efficacy of Wolbachia-Infected Mosquito Deployments for the Control of Dengue. New England Journal of Medicine 384, 2177–2186 (2021). URL http://www.nejm.org/doi/10.1056/NEJMoa2030243.

[15] Tricou, V., et al. Long-term efficacy and safety of a tetravalent dengue vaccine (TAK-003): 4·5-year results from a phase 3, randomised, double-blind, placebo-controlled trial. The Lancet Global Health 12, e257–e270 (2024). URL https://linkinghub.elsevier.com/retrieve/pii/S2214109X23005223.

[16] Dalziel, B. D. et al. Urbanization and humidity shape the intensity of influenza epidemics in U.S. cities. Science 362, 75–79 (2018). URL https://www.science.org/doi/10.1126/science.aat6030.

[17] Ribeiro Dos Santos, G., et al. Estimating the effect of the wMel release programme on the incidence of dengue and chikungunya in Rio de Janeiro, Brazil: a spatiotemporal modelling study. The Lancet Infectious Diseases 22, 1587–1595 (2022). URL https://linkinghub.elsevier.com/retrieve/pii/S1473309922004364.

[18] Pandey, A. & Medlock, J. The introduction of dengue vaccine may temporarily cause large spikes in prevalence. Epidemiology and Infection 143, 1276–1286 (2015).

[19] Okamoto, K. W., Gould, F. & Lloyd, A. L. Integrating Transgenic Vector Manipulation with Clinical Interventions to Manage Vector-Borne Diseases. PLoS computational biology 12, e1004695 (2016).

[20] Hollingsworth, B., Okamoto, K. W. & Lloyd, A. L. After the honeymoon, the divorce: Unexpected outcomes of disease control measures against endemic infections. PLOS Computational Biology 16, e1008292 (2020). URL https://dx.plos.org/10.1371/journal.pcbi.1008292.

[21] Guillen-Calle, B. E., Barja-Ore, J., Roman-Lazarte, V., Trujillo-Sanchez, K. G. & Tello-García, M. Public Perception of Wolbachia-based Dengue Control in High-incidence Countries: A Scoping Review. Journal of South Asian Federation of Obstetrics and Gynaecology 17, S209–S217 (2025). URL https://www.jsafog.com/doi/10.5005/jp-journals-10006-2618.

[22] Pinto, J. et al. Susceptibility to insecticides and resistance mechanisms in three populations of Aedes aegypti from Peru. Parasites & Vectors 12, 494 (2019). URL https://parasitesandvectors.biomedcentral.com/articles/10.1186/s13071-019-3739-6.

[23] Dorigatti, I., McCormack, C., Nedjati-Gilani, G. & Ferguson, N. M. Using Wolbachia for Dengue Control: Insights from Modelling. Trends in Parasitology 34, 102–113 (2018). URL https://linkinghub.elsevier.com/retrieve/pii/S1471492217302726.

[24] Ooi, E.-E., Goh, K.-T. & Gubler, D. J. Dengue prevention and 35 years of vector control in Singapore. Emerging Infectious Diseases 12, 887–893 (2006).

[25] WHO position paper on dengue vaccines - May 2024. Weekly epidemiological record 99, 203–224 (2024). URL https://www.who.int/publications/i/item/who-wer-9918-203-224.

[26] Lim, A. et al. The overlapping global distribution of dengue, chikungunya, Zika and yellow fever. Nature Communications 16, 3418 (2025). URL https://www.nature.com/articles/s41467-025-58609-5.

[27] O’Reilly, K. M. et al. Estimating the burden of dengue and the impact of release of wMel Wolbachia-infected mosquitoes in Indonesia: a modelling study. BMC Medicine 17, 172 (2019). URL https://bmcmedicine.biomedcentral.com/articles/10.1186/s12916-019-1396-4.

[28] A review of transmission models of dengue: A quantitative and qualitative analysis of model features. In Gubler, D. J., Ooi, E. E., Vasudevan, S. & Farrar, J. (eds.) Dengue and dengue hemorrhagic fever (CABI, UK, 2014), 2 edn. URL http://www.cabidigitallibrary.org/doi/book/10.1079/9781845939649.0000.

[29] Reich, N. G. et al. Interactions between serotypes of dengue highlight epidemiological impact of cross-immunity. Journal of The Royal Society Interface 10, 20130414 (2013). URL https://royalsocietypublishing.org/doi/10.1098/rsif.2013.0414.

[30] Tian, Y. et al. Rising dengue risk with increasing El Niño-Southern Oscillation amplitude and teleconnections. Nature Communications 16, 8629 (2025).

[31] Alkuzweny, M., España, G. & Perkins, T. A. Refining uncertainty about the TAK-003 dengue vaccine with a multi-level model of clinical efficacy trial data (2025). URL https://elifesciences.org/reviewed-preprints/108316v1.

[32] Gerardin, J. & Penny, M. A. How can modeling responsibly inform decision-making in malaria? PLOS Biology 23, e3002991 (2025). URL https://dx.plos.org/10.1371/journal.pbio.3002991.

[33] Brady, O. J. et al. The cost-effectiveness of controlling dengue in Indonesia using wMel Wolbachia released at scale: a modelling study. BMC Medicine 18, 186 (2020). URL https://bmcmedicine.biomedcentral.com/articles/10.1186/s12916-020-01638-2.

[34] Smith, D. L., et al. Ross, Macdonald, and a Theory for the Dynamics and Control of Mosquito-Transmitted Pathogens. PLoS Pathogens 8, e1002588 (2012). URL https://dx.plos.org/10.1371/journal.ppat.1002588.

[35] El Hindi, T., et al. Estimated Efficacy of TAK-003 Against Asymptomatic Dengue Infection in Children and Adolescents Participating in the DEN-301 Trial in Asia Pacific and Latin America. The Journal of Infectious Diseases 231, e1160–e1169 (2025). URL https://academic.oup.com/jid/article/231/6/e1160/8084909.

[36] World Population Prospects. URL https://population.un.org/wpp/.

[37] C3S. ERA5 hourly data on single levels from 1940 to present (2018). URL https://cds.climate.copernicus.eu/doi/10.24381/cds.adbb2d47.

[38] Bogoch, I. I. et al. Potential for Zika virus introduction and transmission in resource-limited countries in Africa and the Asia-Pacific region: a modelling study. The Lancet Infectious Diseases 16, 1237–1245 (2016). URL https://linkinghub.elsevier.com/retrieve/pii/S1473309916302705.

[39] Shepard, D. S., Undurraga, E. A., Halasa, Y. A. & Stanaway, J. D. The global economic burden of dengue: a systematic analysis. The Lancet Infectious Diseases 16, 935–941 (2016). URL https://linkinghub.elsevier.com/retrieve/pii/S1473309916001468.

[40] Clarke, J. et al. A global dataset of publicly available dengue case count data. Scientific Data 11, 296 (2024).

[41] Clarke, J., et al. OpenDengue: data from the OpenDengue database (2025). URL https://figshare.com/articles/dataset/OpenDengue_V1_2/24259573. Artwork Size: 109884104 Bytes Pages: 109884104 Bytes.

[42] Lenhart, S. & Workman, J. T. Optimal Control Applied to Biological Models (Chapman and Hall/CRC, 2007), 0 edn. URL https://www.taylorfrancis.com/books/9781420011418.

[43] mrc-ide/arbomap (2024). URL https://github.com/mrc-ide/arbomap. Original-date: 2024-03-18T09:16:12Z.

[44] Wood, S. mgcv: Mixed GAM Computation Vehicle with Automatic Smoothness Estimation (2025). URL https://cran.r-project.org/web/packages/mgcv/index.html.

[45] Chan, M. & Johansson, M. A. The Incubation Periods of Dengue Viruses. PLoS ONE 7, e50972 (2012). URL https://dx.plos.org/10.1371/journal.pone.0050972.

[46] Lambrechts, L. et al. Impact of daily temperature fluctuations on dengue virus transmission by *Aedes aegypti*. Proceedings of the National Academy of Sciences 108, 7460–7465 (2011). URL https://pnas.org/doi/full/10.1073/pnas.1101377108.

[47] Perkins, T. A., Siraj, A. S., Ruktanonchai, C. W., Kraemer, M. U. G. & Tatem, A. J. Model-based projections of Zika virus infections in childbearing women in the Americas. Nature Microbiology 1, 16126 (2016). URL https://www.nature.com/articles/nmicrobiol2016126.

[48] Cavany, S. M., et al. Fusing an agent-based model of mosquito population dynamics with a statistical reconstruction of spatio-temporal abundance patterns. PLOS Computational Biology 19, e1010424 (2023). URL https://dx.plos.org/10.1371/journal.pcbi.1010424.

[49] Gunning, C. E. et al. Efficacy of Aedes aegypti control by indoor Ultra Low Volume (ULV) insecticide spraying in Iquitos, Peru. PLoS neglected tropical diseases 12, e0006378 (2018).

[50] Tiley, K., Entwistle, J., Thomas, B., Yakob, L. & Brady, O. Using models and maps to inform Target Product Profiles and Preferred Product Characteristics: the example of Wolbachia replacement. Gates Open Research 7, 68 (2024). URL https://gatesopenresearch.org/articles/7-68/v3.

[51] Cavany, S., et al. Does ignoring transmission dynamics lead to underestimation of the impact of interventions against mosquito-borne disease? BMJ Global Health 8, e012169 (2023). URL https://gh.bmj.com/lookup/doi/10.1136/bmjgh-2023-012169.

[52] Ferguson, N. M., et al. Modeling the impact on virus transmission of *Wolbachia* - mediated blocking of dengue virus infection of *Aedes aegypti*. Science Translational Medicine 7 (2015). URL https://www.science.org/doi/10.1126/scitranslmed.3010370.

[53] Barbosa, A. D. M. & Veronezi, R. J. B. Dengue control in the state of goias-brazil using “wmel wolbachia”: a cost-effectiveness study. Revista Científica da Escola Estadual de Saúde Pública de Goiás *Cândido Santiago* 9 (2023). URL https://www.revista.esap.go.gov.br/index.php/resap/article/view/672/340.

[54] Pinto, L. How billions of hacked mosquitoes and a vaccine could beat the deadly dengue virus. Nature 645, 578–580 (2025).

