## Supplemental Information for "Combining new interventions with urban development as a path to effective, consistent, and durable control of dengue"

### Supplemental Methods

#### Model description

##### Baseline transmission dynamics

The state variables for the model consist of susceptible ($S$), infectious ($I$), and recovered ($R$) compartments, distinguished by subscripts for the number of previous DENV serotype exposures ($j$). Individuals in $R$ compartments were recently infected, are no longer infectious, and are protected against infection by any serotype due to temporary cross immunity. Once the period of temporary cross immunity passes, they move from $R_j$ to $S_j$ to await their next serotype infection. In total, there are twelve state variables that individuals pass through in the following sequence: $S_0$, $I_1$, $R_1$, $S_1$, $I_2$, $R_2$, $S_2$, $I_3$, $R_3$, $S_3$, $I_4$, and $R_4$. There is also one state variable ($C$) that tracks the proportion of the population living in areas where enhanced vector control is in effect at time $t$.

The dynamics of individuals who are susceptible to all four DENV serotypes follow

$$\frac{dS_0}{dt} = -\Lambda(t)S_0 - \delta_0 S_0 + \upsilon S_0 + \mu N \tag{1}$$

where the first term on the right-hand side captures infection of susceptibles at rate $\Lambda(t)$, the second term results in mortality at rate $\delta_0(t)$, the third term accounts for immigration into the population at per-capita rate $\upsilon$, and the fourth term replenishes the $S_0$ compartment with births at per-capita rate $\mu(t)$. The variable $N$ represents the sum of all $S$, $I$, and $R$ state variables.

The dynamics of individuals who have experienced $j$ serotype infections and remain susceptible to $4 - j$ other serotypes follow

$$\frac{dS_j}{dt} = -\Lambda(t)\frac{4-j}{4}S_j - \delta_j S_j + \upsilon S_j + \eta R_j \tag{2}$$

which are subject to a reduction in the force of infection by a factor $\frac{4-j}{4}$. This assumes that the four DENV serotypes circulate evenly. Otherwise, Eqn. (2) is similar to Eqn. (1), except that the former lacks an input from births but gains an input from individuals from $R_{j-1}$ as their temporary cross-immunity is lost at rate $\eta$. We also note here that the death rate differs for each group $j$, given that each such group is composed of individuals of different ages.

The dynamics of infectious individuals who have experienced $j$ serotype infections follow

$$\frac{dI_j}{dt} = \Lambda(t)\frac{4-j+1}{4}S_{j-1} - \delta_j I_j + \upsilon I_j - \gamma I_j, \tag{3}$$

which accounts for a reduction in the force of infection by a factor $\frac{4-j+1}{4}$ given that susceptibles who transition into $I_j$ have only experienced $j - 1$ serotype infections. In addition to the other transitions to which other state variables are subject, individuals in the $I$

compartments transition into $R$ compartments at rate $\gamma$, which is the rate of recovery from infectiousness.

The last remaining compartment follows

$$\frac{dR_j}{dt} = -\delta_j R_j + \gamma I_j + \upsilon R_j - \eta R_j, \tag{4}$$

where $R_j$ includes individuals who have experienced $j$ serotype infections and still possess temporary cross immunity. When $j = 4$, there is no term involving $\eta$ because individuals are permanently immune to all serotypes after having experienced all four already.

The rate $\Lambda(t)$, also known as the force of infection, sums the rate of infection from all $I_j$ compartments according to

$$\Lambda(t) = \Lambda_i + \beta(t) \sum_{i=1}^{4} \frac{I_j}{N}, \tag{5}$$

which assumes that individuals are subject to infection through importation of the virus from elsewhere, which occurs at rate $\Lambda_i$ and helps prevent the $I_j$ variables from reaching values that are numerically indistinguishable from zero during the model calibration. The function $\beta(t)$ represents the time-varying transmission coefficient in areas untreated with enhanced vector control, taking the form

$$\beta(t) = \frac{\epsilon(t) a^2(t) b(t) c(t) e^{-g(t)n(t)}}{g^2(t)} \tag{6}$$

after the Ross-Macdonald model of mosquito-borne pathogen transmission [34]. All component functions are time-varying and differ for each city as a function of its environmental conditions.

#### Transmission dynamics under control

We assume that enhanced vector control temporarily reduces mosquito densities to a proportion $\phi_c$ of their baseline value in areas where it is in effect. If we assume that enhanced vector control is applied at a time-varying rate $u_c(t)$ up to a maximum coverage $C_{c,\max}$ and that its effectiveness wanes at rate $\rho_c$, then we can describe the dynamics of its coverage with

$$\frac{dC_c}{dt} = u_c(t) \left(1 - \frac{C_c}{C_{c,\max}}\right) - \rho_c C_c. \tag{7}$$

Relative to a model without enhanced vector control, the transmission coefficient becomes

$$\beta_c(t) = (C_c(t)\phi_c + (1 - C_c(t)))\beta(t), \tag{8}$$

which assumes proportionate mixing between areas with and without enhanced vector control. We assume that *Wolbachia* permanently reduces mosquito infectiousness to a proportion $\phi_w$ of its baseline value in areas where it is in effect. We assume that *Wolbachia* coverage increases linearly over a five-year period to its maximum, such that

$$C_w(t) = \begin{cases} \frac{t}{5} C_{w,\max}, & t < 5 \\ C_{w,\max}, & t \geq 5 \end{cases}, \tag{9}$$

Relative to a model without *Wolbachia*, the transmission coefficient becomes

$$\beta_w(t) = (C_w(t)\phi_w + (1 - C_w(t)))\beta(t). \tag{10}$$

Building out has a similar effect on the model as *Wolbachia*, except that its coverage increases linearly over 25 years and its effect on transmission, captured by $\phi_b$, is conceptualized as reducing mosquito density. For situations in which two or more of enhanced vector control, *Wolbachia*, or building out are in effect, their effects on $\beta(t)$ combine multiplicatively. Because we assume that vaccination has a negligible effect on blocking infection, it has no impact on $\beta(t)$ and instead affects disease outcomes and associated costs.

#### Costs

We calculated the aggregate costs of disease and control

$$J = J_D + J_C \tag{11}$$

over a time frame of 25 years, where $J_D$ are costs associated with disease and $J_C$ are costs associated with control. In model scenarios without vaccination, the cost of disease is

$$J_D = \int_0^{25} \left( \sum_o w_o \sum_{j=0}^{3} p_o^j I_j(t) \right) dt, \tag{12}$$

where $w_o$ is the cost associated with an infection resulting in disease outcome $o$ and $p_o^j$ is the probability that a new infection results in disease outcome $o$ in someone with $j$ previous infections. The possible disease outcomes are $n$ (non-medical), $a$ (ambulatory), $h$ (hospitalized), and $f$ (fatal), after Shepard et al. [39]. In model scenarios with vaccination, the cost of disease is modified to

$$J_D = \int_0^{25} \left( \sum_o w_o \sum_{j=0}^{3} \left( V(t) p_o^{j,v} + (1 - V(t)) p_o^j \right) I_j(t) \right) dt, \tag{13}$$

where $V(t)$ is vaccination coverage and

$$p_n^{j,v} = p_n^j + (1 - RR_a^j) p_a^j \tag{14}$$

$$p_a^{j,v} = RR_a^j p_a^j + (1 - RR_{hos}^j)(p_h^j + p_f^j) \tag{15}$$

$$p_h^{j,v} = RR_h^j p_h^j \tag{16}$$

$$p_f^{j,v} = RR_h^j p_f^j. \tag{17}$$

These modified probabilities, $p^v_{j,o}$, reflect the amelioration of ambulatory disease and hospitalization by their respective relative risks, $RR^j_a$ and $RR^j_h$. $V(t)$ is set such that target vaccination coverage, $V_{\max}$, is achieved among individuals aged 6-16 years-old within the first year of vaccination and among new 6 year-olds in each year thereafter.

The cost of control is

$$J_C = J_c + J_w + J_v + J_b, \tag{18}$$

which is the sum of the costs of each of the four interventions. The cost of enhanced vector control

$$J_c = \int_0^{25} w_c u_c(t) dt \tag{19}$$

is proportional to the extent of its deployment, with $w_c$ being the per-person cost associated with one-time application of enhanced vector control. The cost of *Wolbachia* is envisioned as a one-time, up-front cost per person,

$$J_w = w_w. \tag{20}$$

The cost of vaccination is

$$J_v = \sum_{t=1}^{25} w_v(V(t) - V(t-1)), \tag{21}$$

where $w_v$ is the cost of fully vaccinating a single individual with the required two doses. In light of the many avenues by which building out could be implemented in different settings, we do not attempt to specify a value for this quantity.

#### Optimal control

Whereas the coverage of *Wolbachia*, vaccination, and building out are all pre-specified, we chose to identify appropriate coverages of enhanced vector control, $C_c(t)$, using optimal control theory. To do this, we used an approach described by Lenhart and Workman [42] to identify values of the time-varying rate of deployment of enhanced vector control, $u_c(t)$, that minimized the overall cost $J$ from time $t = 0$ to 25.

Identifying the optimal control first required defining the Hamiltonian,

$$
\begin{aligned}
\mathcal{H} = J \,+\, & \lambda_{S_0}\left(-\Lambda S_0 - \delta_0 S_0 + \upsilon S_0 + \mu N\right) \\
+ & \sum_{j=1}^{3} \lambda_{S_j}\left(-\Lambda \frac{4-j}{4} S_j - \delta_j S_j + \upsilon S_j + \eta R_j\right) \\
+ & \sum_{j=1}^{4} \lambda_{I_j}\left(\Lambda \frac{4-j+1}{4} S_{j-1} - \delta_j I_j + \upsilon I_j - \gamma I_j\right) \\
+ & \sum_{j=1}^{3} \lambda_{R_j}\left(-\delta_j R_j + \gamma I_j + \upsilon R_j - \eta R_j\right) \\
+ & \lambda_{R_4}\left(-\delta_4 R_4 + \gamma I_4 + \upsilon R_4\right) \\
+ & \lambda_C\left(u_c\left(1 - \frac{C_c}{C_{c,max}}\right) - \rho_c C_c\right)
\end{aligned} \tag{22}
$$

Next, we derived the adjoint equations by applying Pontryagin's maximum principle and defining the differential equation of the adjoint variable $\lambda_x$ as

$$\lambda_x t = -Hx. \tag{23}$$

The series of adjoint equations then becomes

$$
\begin{aligned}
\lambda_{S_0} t = {} & \lambda_{S_0}\left(\Lambda + \delta_0 - \mu - \upsilon\right) + \sum_{k=0}^{3} \lambda_{S_k}\left(-\frac{4-k}{4}\frac{S_k}{N}(\Lambda - \Lambda_0) - \frac{S_k}{N}(\delta_0 - \mu - \upsilon)\right) \\
& + \sum_{k=1}^{4} \lambda_{I_k}\left(\frac{4-k+1}{4}\frac{S_{k-1}}{N}(\Lambda - \Lambda_0) - \frac{I_k}{N}(\delta_0 - \mu - \upsilon)\right) + \lambda_{I_1}(-\Lambda) + \sum_{k=1}^{4} \lambda_{R_k}\left(-\frac{R_k}{N}(\delta_0 - \mu - \upsilon)\right)
\end{aligned} \tag{24}
$$

$$
\begin{aligned}
\lambda_{S_i} t = {} & \sum_{k=0}^{3} \lambda_{S_k}\left(-\frac{4-k}{4}\frac{S_k}{N}(\Lambda - \Lambda_0) - \frac{S_k}{N}(\delta_i - \mu - \upsilon)\right) + \lambda_{S_i}\left(\frac{4-i}{4}\Lambda + \delta_i - \upsilon\right) \\
& + \sum_{k=1}^{4} \lambda_{I_k}\left(\frac{4-k+1}{4}\frac{S_{k-1}}{N}(\Lambda - \Lambda_0) - \frac{I_k}{N}(\delta_i - \mu - \upsilon)\right) - \frac{4-i}{4}\Lambda \lambda_{I_{i+1}} \\
& + \sum_{j=1}^{4} \lambda_{Rj}\left(-\frac{R_j}{N}(\delta_i - \mu - \upsilon)\right) \text{ for } i = 1, 2, 3,
\end{aligned} \tag{25}
$$

$$\lambda_{I_i}t = \sum_{k=0}^{3} \lambda_{S_k} \left( \frac{4-k}{4} S_k(\Lambda - \Lambda_0) \left( \frac{1}{I} - \frac{1}{N} \right) - \frac{S_k}{N}(\delta_i - \mu - \upsilon) \right) + \lambda_{I_i} \left( \delta_i + \gamma - \upsilon \right)$$
$$+ \sum_{k=1}^{4} \lambda_{I_k} \left( -\frac{4-k+1}{4} S_{k-1}(\Lambda - \Lambda_0) \left( \frac{1}{I} - \frac{1}{N} \right) - \frac{I_k}{N}(\delta_i - \mu - \upsilon) \right) + \lambda_{R_i} \left( -\gamma \right) \quad (26)$$
$$+ \sum_{j=1}^{4} \lambda_{Rj} \left( -\frac{R_j}{N}(\delta_i - \mu - \upsilon) \right) - w_{I_i}\gamma \text{for } i = 1,2,3,4,$$

$$\lambda_{R_i}t = \sum_{k=0}^{3} \lambda_{S_k} \left( -\frac{4-k}{4} \frac{S_k}{N}(\Lambda - \Lambda_0) - \frac{S_k}{N}(\delta_i - \mu - \upsilon) \right) + \lambda_{S0} \left( -\mu \right) + \mathbf{1}_{\{i \le 3\}} \lambda_{S_i}(-\eta)$$
$$+ \sum_{k=1}^{4} \lambda_{I_k} \left( \frac{4-k+1}{4} \frac{S_{k-1}}{N}(\Lambda - \Lambda_0) - \frac{I_k}{N}(\delta_i - \mu - \upsilon) \right) + \sum_{j=1}^{4} \lambda_{R_j} \left( -\frac{R_j}{N}(\delta_i - \mu - \upsilon) \right)$$
$$+ \lambda_{R_i} \left( \delta_i + \mathbf{1}_{\{i \le 3\}}\eta - \upsilon \right) \text{ for } i = 1,2,3,4, \text{ and} \quad (27)$$

$$\lambda_C t = w_c \frac{u_c}{C_{c,\max}} + \sum_{k=0}^{3} \lambda_{S_k} \left( \Lambda C_c \frac{4-k}{4} S_k \right) + \sum_{k=1}^{4} \lambda_{I_k} \left( -\Lambda C_c \frac{4-k+1}{4} S_{k-1} \right) + \lambda_C \left( \frac{u_c}{C_{c,\max}} + \rho \right), \quad (28)$$

where

$$\begin{aligned}
\Lambda C_c &= (\beta_c - \beta_n) \sum_{j=1}^{4} \frac{I_j}{N}, \\
I &= I_1 + I_2 + I_3 + I_4, \\
N &= S_0 + S_1 + S_2 + S_3 + I_1 + I_2 + I_3 + I_4 + R_1 + R_2 + R_3 + R_4, \\
\Lambda &= \Lambda_0 + \beta_c C_c \frac{I}{N} + \beta_n (1 - C_c) \frac{I}{N}, \\
\delta &= \delta_0 \frac{S_0}{N} + \delta_1 \frac{I_1 + R_1 + S_1}{N} + \delta_2 \frac{I_2 + R_2 + S_2}{N} + \delta_3 \frac{I_3 + R_3 + S_3}{N} + \delta_4 \frac{I_4 + R_4}{N}, \\
\upsilon &= -\mu + \delta,
\end{aligned} \quad (29)$$

and $\mathbf{1}_{\{i \le 3\}}$ denotes the indicator function, equal to 1 if $i \le 3$ and 0 otherwise.

The optimal control $u_c^*$ that minimizes the objective functional $J$ is determined by the switching function

$$\psi = \frac{\partial \mathcal{H}}{\partial u_c} = w_{SR} \left( 1 - \frac{C_c}{C_{c,\max}} \right) + \lambda_C \left( 1 - \frac{C_c}{C_{c,\max}} \right). \quad (30)$$

To investigate whether the control problem is bang-bang and/or singular, suppose $u_c^*$ is singular on some interval, say $(0, l)$. Then, on this interval, $\psi = 0$ so that

$$\lambda_C \left(1 - \frac{C_c}{C_{c,\max}}\right) = -w_{SR} \left(1 - \frac{C_c}{C_{c,\max}}\right). \tag{31}$$

Differentiating both sides of (31) yields

$$\left(1 - \frac{C_c}{C_{c,\max}}\right) \frac{d\lambda_C}{dt} - \frac{\lambda_C}{C_{c,\max}} \frac{dC_c}{dt} - \frac{w_{SR}}{C_{c,\max}} \frac{dC_c}{dt} = 0, \tag{32}$$

which is impossible based on the formulations of $\frac{d\lambda_c}{dt}$ and $\frac{dC_c}{dt}$. Hence, we conclude that $u_c^*$ is nowhere singular and therefore the control problem is bang-bang. Consequently, the optimal control is given by

$$u_c^* = \begin{cases} u_{\max,c}, & \text{if } \psi < 0, \\ 0, & \text{if } \psi > 0. \end{cases} \tag{33}$$

After obtaining all required equations, we use the forward-backward sweep method (FBSM) to solve the system of state variables and the system of adjoint variables [42]. For this purpose, we set the state variables to zero at the initial time (initial conditions) and adjoint variables to zero at the final time (transversality conditions).

#### Model parameterization

##### Geographic inputs

We used a data set on cities from simplemaps [36], which includes 48,000 locations globally. Relevant fields from this data set include the latitude, longitude, administrative units, and population of each city. Consistent with our interest in urban areas in settings where DENV circulates, we subsetted this data set to locations with populations of 100,000 or more and DENV force of infection of 0.005 or greater [11]. This resulted in a total of 1,634 cities spread across 76 countries. The specific version of DENV force of infection estimates that we used in our analysis come from ArboMap [43], which provides updated estimates of DENV force of infection using the same methods as [11] but with the addition of new data that has become available since then.

We based temperature inputs for transmission parameters on ERA5 reanalysis projections [37]. We used the "2m temperature" variable (available at 10-by-10 minute resolution) averaged monthly from 2015 to 2024 for 3:00 am and 3:00 pm to separately capture daily minimum and maximum temperatures. We then used the approxfun function in R to establish a function capable of generating continuous subdaily temperatures based on linear interpolation between the daily minimum and maximum temperatures.

To inform seasonal patterns of mosquito density, which could be influenced by factors other than temperature (e.g., precipitation, urbanization), we worked with monthly estimates of *Ae. aegypti* occurrence probabilities at an approximately 5 km by 5 km resolution globally [38]. To interpolate these monthly values to daily values, we fitted a generalized additive model to each set of 12 monthly occurrence probabilities from each city using the mgcv package in R [44]. We used cyclic cubic regression splines to ensure that the beginning and end of the year had the same value, and we specified the number of knots to be 5 to achieve a balance between flexibility and overfitting.

#### Transmission parameters

The time-varying transmission coefficient included five temperature-dependent parameters: $a$, blood-feeding rate; $b$, mosquito infectiousness; $c$, mosquito susceptibility; $g$, adult mosquito mortality; $n$, incubation period in the mosquito. All relationships between temperature and these parameters were informed by laboratory experiments, with relationships for $a$, $b$, $c$, and $g$ obtained from [13] and $n$ obtained from [45]. Some of these relationships use functional forms that take on values of zero or less at unsuitable temperatures, which leads to numerical difficulties with solving the model. To make the model robust to a wide range of temperature inputs while still preserving the tendency for unsuitable temperatures to lead to the cessation of transmission, we approximated temperature relationships with this property using skewed generalized t density functions fitted by least squares. These functions had sufficient flexibility to approximate previously estimated temperature relationships without taking on values of zero or less. We computed values of each of the five aforementioned parameters on a sub-daily basis to account for diurnal temperature variation, which we collapsed to daily values for use in the model via rate summation [46].

To estimate the time-varying mosquito emergence rate, $\epsilon(t)$, we took monthly mosquito occurrence probabilities, $OP_t$, smoothed them with a generalized additive model using the gam package in R to obtain $OP(t)$, and converted them to relative mosquito densities, $\tilde{m}(t)$, according to the relationship $\tilde{m} = -\ln(1 - OP)$ [47]. If mosquitoes emerge at rate $\epsilon$ and live for $1/g$ days, then $\tilde{m} = \tilde{\epsilon}/g$ and $\tilde{\epsilon}(t) = \tilde{m}(t)g(t)$. To obtain an absolute—and not just relative—emergence rate $\epsilon(t)$, we multiplied $\tilde{\epsilon}(t)$ by $\epsilon^*$, where the latter was tuned so that the time-averaged value of $\Lambda(t)$ equaled the force of infection estimated by Cattarino et al. [11] for a given city.

Other parameters in the transmission model took on assumed values that were held constant, including $\Lambda_i$ (rate of $10^{-5}$ imported infections per person per year), $\gamma$ (average infection recovery of 7 days), and $\eta$ (average cross-immunity of 2 years).

#### Demographic parameters

The human birth rate $\mu$ was taken as the average crude birth rate during 2015-2024 from the country in which a city is located (UN WPP). To calculate the human death rates $\mu_j$, we first obtained average age-specific death rates, $\mu_a$, during 2015-2024 from the country in which a city is located (UN WPP). Next, we used a catalytic model to calculate the proportion of individuals of age $a$ with $j$ previous serotype infections, $p_{a,j}$. Like our dynamic model, this catalytic model allowed for up to four serotype infections and used the same force of infection and duration of temporary cross-immunity. The death rates $\mu_j$ were then calculated as $\sum_a \mu_a p_{a,j} / \sum_a p_{a,j}$. Finally, we set the $v$ parameter to equal the difference between the overall birth and death rates. This resulted in the maintenance of a constant population size due to what can be conceptualized as immigration or emigration irrespective of current or past DENV infection status.

#### Intervention parameters

Our assumptions about the effect of enhanced vector control were designed to match the behavior of an agent-based model of *Aedes aegypti* population dynamics [48] informed by empirical observations of ultra-low volume indoor adulticide spraying in Iquitos, Peru [49]. Based on that information, we set $\phi_{\epsilon,c} = 0.032$, $\rho_c = 14.54$ yr$^{-1}$, $u_{\max,c} = 13.52$ yr$^{-1}$, and $C_{\max,c} = 0.62$. Roughly speaking, those parameter values mean that spraying kills 97% of mosquitoes in treated houses, that the mosquito densities in those houses rebound in 25 days on average, and that it takes 27 days to achieve the empirically observed coverage of 62% of houses in the city. We modeled the cost of enhanced vector control based on a systematic review of vector control costing studies, which estimated a current routine cost of vector control in Brazil of $2.03 USD per person per year [50]. In this review, Tiley et al. also noted that monthly routine costs of vector control program increased by approximately 50% during outbreaks, when enhanced vector control is used. This would give an additional cost of conducting enhanced vector control of 0.5 x $2.03/12 = $0.0846 USD per person per month, which is the cost that we assumed for a single application of enhanced vector control.

Our assumptions about the effect of *Wolbachia* follow a modelling study [51] of *wMel Wolbachia* that translated the 77% protective efficacy estimated in a clinical trial [14] into an estimate of the reduction in transmission (50%) that could have generated that efficacy. As others have noted [52], such a reduction in transmission could be achieved through multiple mechanisms, including a reduction in mosquito susceptibility or infectiousness or a lengthening of the extrinsic incubation period. We considered a range of *Wolbachia* penetrance into the mosquito population from 30% to 70%, consistent with wide variation observed across contexts where it has been deployed to date. We model the costs of *Wolbachia* based on a recent cost effectiveness analysis from in the state of Goias, Brazil [53], which estimated an average cost of R$ 34.55 per person, equivalent to approximately $6.50 USD at the time of analysis and approximately in line with previous estimates from other countries after accounting for the reduced costs of larger scale programs [33, 50]. As in previous analyses [33], we assumed this incurred a one-time cost with no ongoing maintenance costs.

Our assumptions about the effect of vaccination are based on a Bayesian statistical analysis of phase-III clinical trial results for the Qdenga vaccine that yields estimates of serotype- and serostatus-dependent per-exposure protection separately against symptomatic disease and hospitalisation [31]. Given that no empirical data currently support a durable effect of vaccination of infection blocking [35], all protection conferred by the vaccine was assumed to affect infection severity only. Based on reported figures from the first large-scale vaccination program in Brazil with Qdenga [54], we assume a one-time cost per person of $38.00 USD to become vaccinated with two doses.

Our assumptions about the effectiveness of building out were made to achieve a steady reduction in dengue virus force of infection consistent with the roughly 1% annual reduction observed during a period of development and modernisation in Singapore [24].

#### Cost of disease parameters

We adopted a set of four possible disease outcomes, $n$ (non-medical), $a$ (ambulatory), $h$ (hospitalized), and $f$ (fatal), after Shepard et al. [39]. These cost estimates were inflated from their 2013 estimate year to the year 2024 using World Bank GDP deflators [36]. To set the probabilities of these four disease outcomes, $o$, as a function of the number of previous DENV infections, $j$, we adopted values of $p_o^j$ assumed by the Notre Dame model in a previously published multi-model analysis of dengue vaccination impacts (derived from Table B in the Supplemental Material of [7]). We set the costs associated with each of these infection outcomes based on values proposed by Shepard et al. [39], which are based on medical and non-medical costs incurred as a direct result of illness with dengue, time lost due to illness or care, and the economic cost associated with premature death. Because we were interested in variation in intervention impacts owing to epidemiological rather than economic differences, we standardized all costs in the analysis to country-specific values for Brazil as a representative dengue-endemic country with multiple cost study estimates.

### Supplemental Results

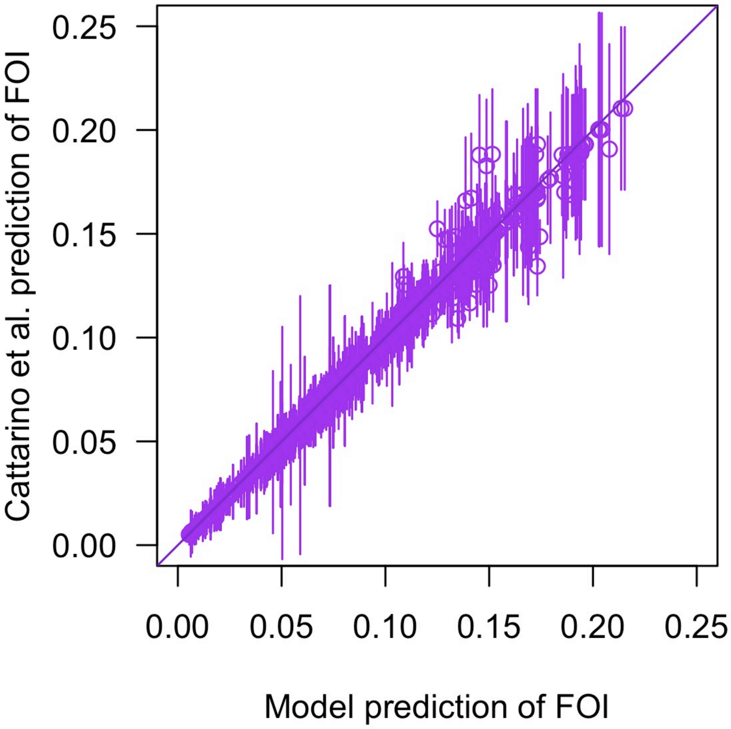


Figure S1: **Comparison of the model's predictions of force of infection against the values to which it was calibrated.** The values of force of infection to which the model was calibrated were generated by Cattarino et al. [11] and are available at https://github.com/mrc-ide/arbomap. In addition to central estimates (circes), intervals one standard deviation above and below the central estimates are also shown (line segments). A one-to-one line is shown in black. The predicted and reference values of force of infection have a Pearson's correlation of 0.997.

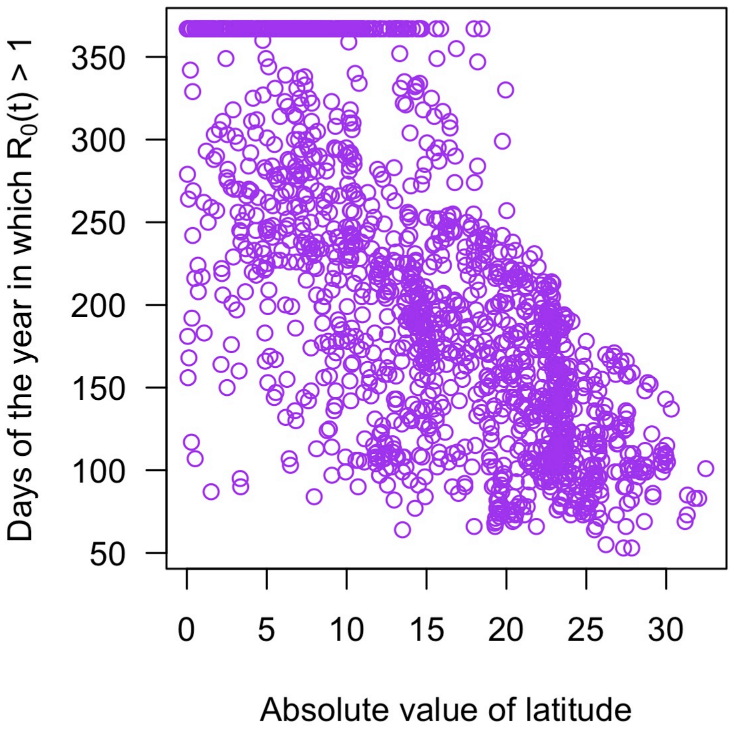


Figure S2: **Days of the year in which $R_0(t) > 1$ against absolute value of latitude.** These variables have a Pearson's correlation of -0.712.

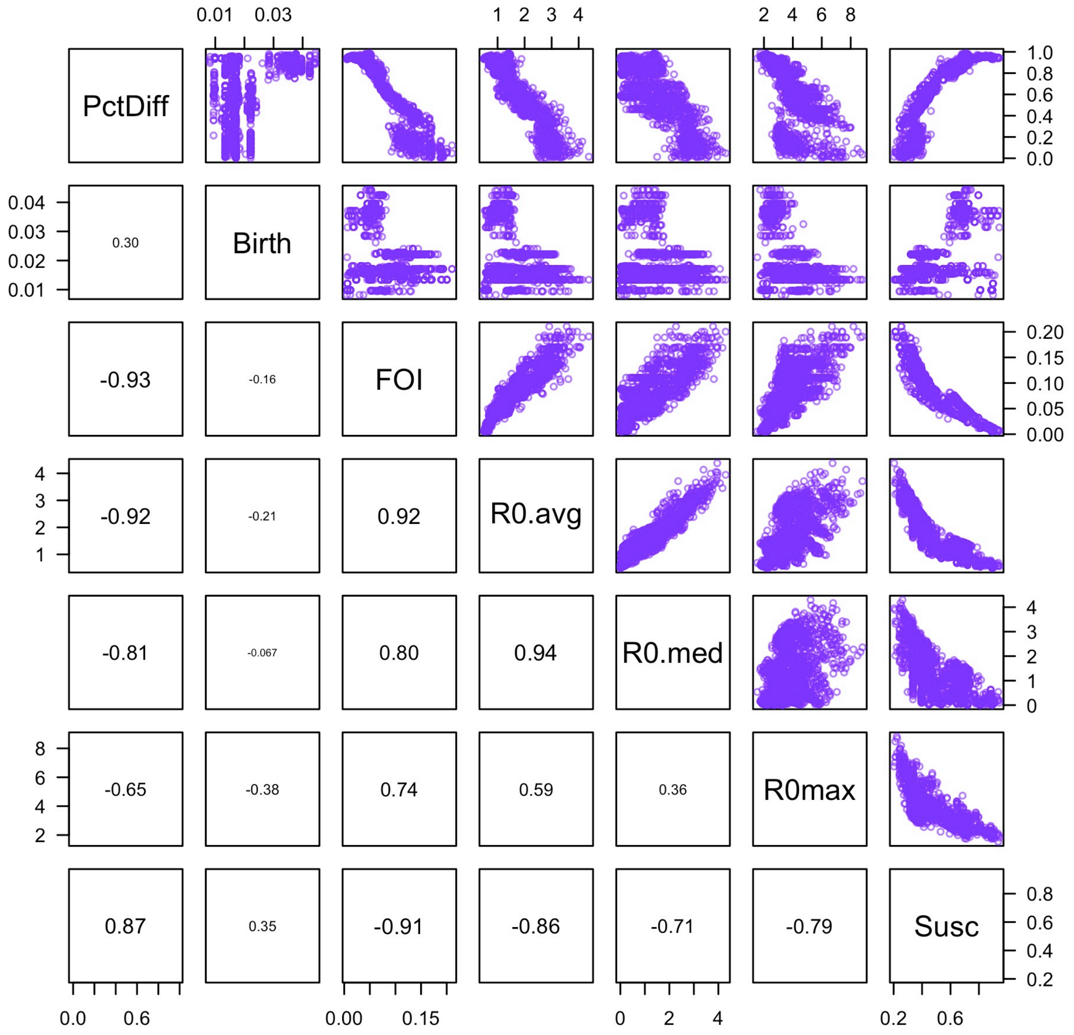


Figure S3: **Pairwise relationships between model-predicted proportional reduction in hospitalisations (PctDiff) and city-level factors.** City-level factors include birth rate (Birth), long-term average force of infection (FOI), seasonal average of $R_0(t)$ (R0.avg), seasonal median of $R_0(t)$ (R0.med), seasonal maximum of $R_0(t)$ (R0.max), and long-term average population susceptibility (Susc). These results correspond to a scenario with enhanced vector control over a 25-year time horizon. Numbers indicate Pearson correlation values.

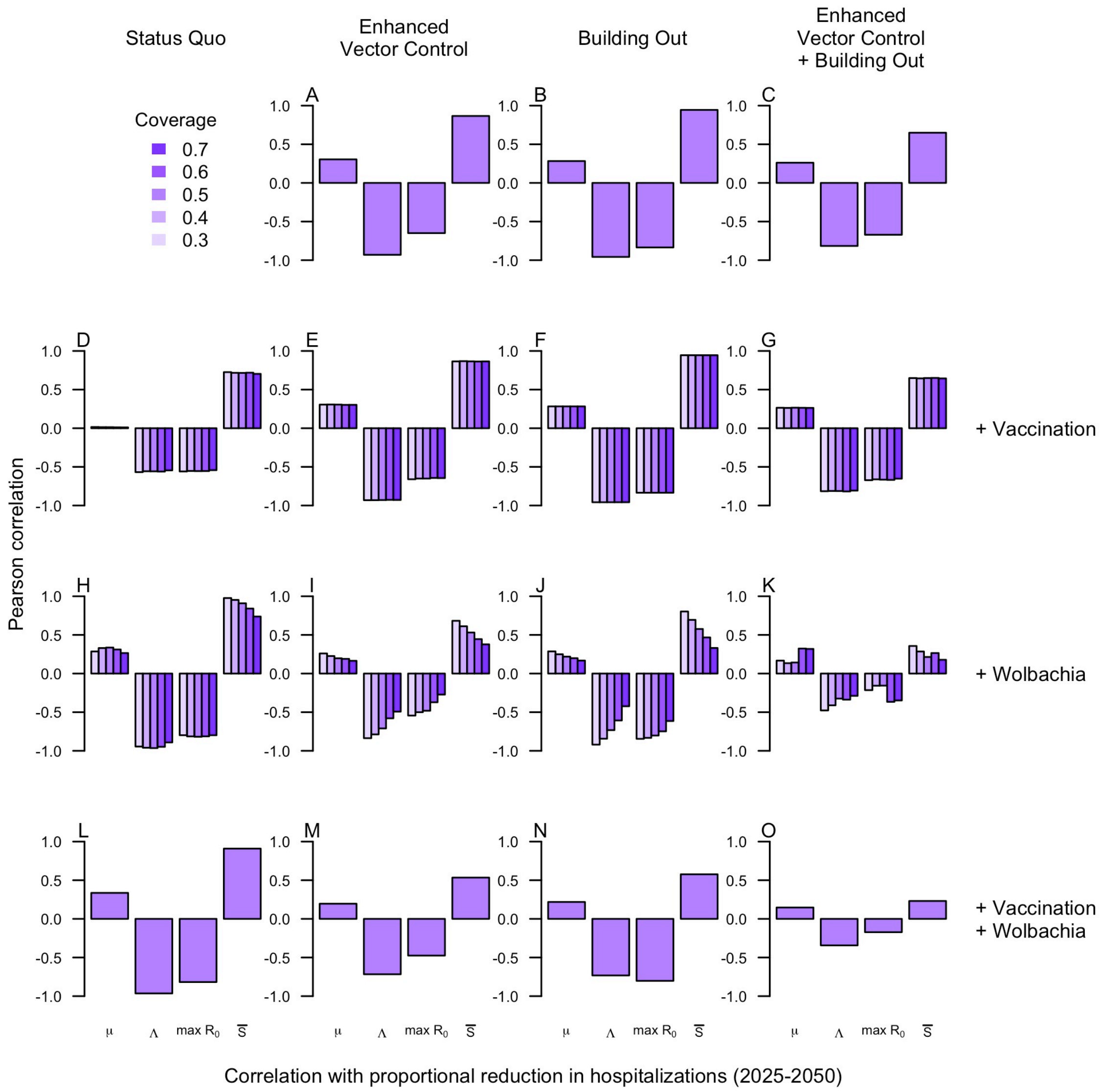


Figure S4: **Pairwise correlations between model-predicted proportional reduction in hospitalisations (y-axis) and city-level factors (x-axis) across intervention scenarios (panels).** City-level factors include birth rate ($\mu$), long-term average force of infection ($\Lambda$), seasonal maximum of $R_0(t)$ ($\max R_0$), and long-term average population susceptibility ($\overline{S}$). These results correspond to a scenario with enhanced vector control over a 25-year time horizon. For vaccination and *Wolbachia*, a range of coverages (colors) were explored. Coverages for enhanced vector control and building out were set at their default values.
